# Access to treatment protocols and manuals for evidence-based psychological interventions for severe mental disorders: A survey of randomized trials included in network meta-analyses

**DOI:** 10.1101/2024.12.26.24319515

**Authors:** Chrysanthi Blithikioti, Giuliano Tomei, Fabrizio Visconti, Lorena Pizzocri, Camilla Cadorin, Irene Gómez Gómez, Ioana Alina Cristea

## Abstract

**Question:** Evidence-based psychological interventions for mental disorders are described in protocols and manuals, but are these resources accessible or publicly available?

**Study selection and analysis:** We surveyed randomized controlled trials (RCTs) from six large network meta-analyses of psychological interventions for severe mental disorders (psychotic, borderline personality, substance use, bipolar, anorexia and bulimia nervosa). Between January 2024 and February 2025, we retrieved protocols and manuals of psychological intervention arms using a multipronged approach (published protocol, trial registries, author contact, commercial availability). We report the proportion of trials and intervention arms for which protocols or manuals were (1) accessible, i.e., retrievable by any method, and (2) publicly versus commercially available.

**Findings:** We included 260 RCTs, with 422 active intervention arms. We retrieved published protocols for 20 RCTs (8%, 95% confidence interval/CI 5% to 12%) and contacted 450 authors for the remaining 240. Authors shared protocols for 43/240 trials (18%, 95% CI 13% to 23%), refused to share for 73 (30%, 95% CI 25% to 37%) and did not respond for 101 (42%, 95% CI 36% to 49%). Protocols or manuals were retrievable for 364 psychological intervention arms (86%, 95% CI 83% to 89%), with 191 available commercially (45%, 95% CI 40% to 50%) and 106 (25%, 95% CI 21% to 30%) publicly.

**Conclusions:** Retrieving detailed descriptions of psychological interventions used in trials, crucial for identifying treatment components, was challenging, resource-intensive and required multiple methods. Reliance on public availability and author sharing enabled access to about 40% of protocols or manuals.

**Funding:** European Union (European Research Council)

**Key messages:** *What is already known on this topic:* Access to treatment protocols or manuals, which detail the component elements of psychological interventions and conditions of application, is paramount to evaluating these treatments. There is no previous systematic overview of the accessibility of protocols and manuals for mental disorders.

*What this study adds:* In a large-scale evaluation of 422 evidence-based psychological interventions for severe mental disorders, studied in 260 randomized trials, sourced from 6 large network meta-analyses, we could retrieve a detailed intervention description, operationalized as a protocol or manual, for about 86% of the active interventions. Around 45% of the protocols or manuals were only available commercially, and only around a quarter were publicly available. The retrieval rates are based on a resource-and time-intensive, multipronged approach, through searching multiple sources (papers, trials registries, Google for commercially available manuals) and extensive author contact.

*How this study might affect research, practice or policy:* Developing psychological treatments and evaluating these in randomized trials need to be complemented by a more streamlined and less taxing access to treatment components and other characteristics (e.g., delivery modes, training required), as for labels of approved drugs. To improve dissemination of effective psychological treatments - a global mental health priority, public availability of treatment manuals is crucial. Funders, journals, and trial registries could require sharing of intervention protocols.

## Background

Psychological interventions for mental disorders are complex, including multiple, potentially interacting, components^1^. They are effectively “packages”, composed of an array of elements, such as practices or techniques, spanning behavioural, interpersonal, cognitive and emotional domains^2^. There is currently no systematically developed, comprehensive taxonomy of components of psychological interventions for adult mental disorders. One existent taxonomy (The Behavior Change Techniques taxonomy^3^) specifies the components of health behavior change interventions. These elements are not intended for symptoms of mental disorders, which are often cognitive, such as delusions or rumination. Moreover, the level of granularity at which behavior change techniques are defined make them difficult to apply to psychotherapies, which are often described along broader components, such as cognitive restructuring.

Identifying active ingredients was identified as a stringent research priority in position papers such as *The Institute of Medicine* report on psychosocial interventions^4^ and *The Lancet Psychiatry Commission*^5^ on psychological treatments research. Conversely, a classification of active ingredients has been developed for common mental disorders in youth^6^, based on 322 randomized controlled trials (RCTs) that tested 615 different treatment protocols, distilled in 41 discrete clinical techniques. Without knowledge of active ingredients, psychological interventions are evaluated as a whole or across broad categories (e.g., cognitive behavioral therapy/CBT). This level of analysis is broad and imprecise, significantly curtailing the investigation of variability in treatment response and of potential mechanisms of action. To address this problem, The European Research Council (ERC)-funded *“Disentangling psychological interventions for mental disorders into a taxonomy of active ingredients”*(DECOMPOSE) project (https://cordis.europa.eu/project/id/101042701) aims to dismantle psychological interventions for severe mental disorders into their constituent elements and subsequently integrate these in an unified, cross-disorder taxonomy, with uniform labels and definitions.

A crucial prerequisite for achieving this goal is access to detailed descriptions of the interventions. Trials of psychological interventions are often poorly reported^7^ ^8^, and interventions in particular are cursorily and incompletely described^9^ as “packages” simply referencing the manual. Intervention protocols or manuals are analogous to the package insert or labels for drugs, and access to them is crucial for identifying constituent elements. Systematic evaluations of availability to these resources have been scarce. One small-scale analysis looked at the availability of manuals for 27 trials of psychological interventions set in low- and middle-income countries^10^. No previous research attempted to retrieve treatment protocols or manuals for evidence-based psychological interventions across multiple mental disorders.

## Objective

We report the first large-scale survey using RCTs selected from network meta-analyses (NMAs) of psychological interventions for severe mental disorders. We examine what proportion of protocols or manuals (1) can be retrieved, using a multiplicity of approaches, and (2) are publicly available, a key factor in dissemination.

## Study selection and analysis

### Selection of studies

The methodology was a survey of randomized trials, sampled from network meta-analyses. No new systematic search was conducted to identify the studies. As reporting guidelines for methodological studies are still under development^11^, we used the PRISMA reporting guidelines insofar as possible.

To assemble a large collection of evidence-based psychological interventions, we assembled a large cohort of RCTs, sourced from published NMAs. NMAs combine large collections of trials, with various comparators, thereby ensuring that both frequently and infrequently studied interventions are represented. Included trials are routinely rated for risk of bias, allowing for ancillary analyses restricted to the highest-quality trials. Systematic reviews accompanied by NMAs were proposed as the highest level of evidence in treatment guidelines^12^ and are being increasingly used in evaluating psychological interventions^13^.

To be included, NMAs had to examine the effect of psychological interventions for severe mental disorders, chosen because they received limited attention in terms of identifying and evaluating treatments components. Severe mental disorders were considered as those higher risk of mortality (all-cause or suicide), and generally poorer treatment outcomes compared to more common mental disorders. We included schizophrenia and psychotic, bipolar disorders, eating disorders (bulimia and anorexia nervosa, substance use disorders (SUD) and borderline personality disorder. For each disorder, we queried Pubmed as of November 2023 (S1 Table) for NMAs published over the last 5 years (i.e., November 2018). One NMA was selected for each disorder prioritizing number of included trials, recency (if similar number of trials) and inclusion of psychological interventions as stand-alone, versus adjunctive interventions (when available). Search strings and reasons for exclusion are presented in S1 Table.

From each of the six NMAs^14–19^, we selected all included RCTs and for each RCT, all active psychological intervention arms. These were defined as having psychological components and being delivered within the trial according to a protocol or manual. Interventions that the trialists intended as control were included, provided they fit these conditions, based on the rationale that an intervention initially developed as control is often shown to have therapeutic benefits. For example, general psychiatric management, intended as a control condition, showed similar efficacy with dialectical behavior therapy, intended as the experimental intervention, for borderline personality disorder symptoms^20^. Treatment as usual (TAU) or enhanced TAU interventions were excluded, as these are often not delivered according to a protocol and there is often no verification that all participants receive the same intervention. Active interventions with no psychological components were also excluded.

### Identification and retrieval of treatment protocols

We used a multipronged approach combining various sources of information, to identify intervention protocols or manuals for RCTs and arms. Protocols were defined as detailed descriptions of the intervention, for example by session or by modules, developed for the purpose of the trial. Manuals were defined as the full description of the intervention, usually as a book written by the treatment developer. First, for each trial report, pairs of researchers (CB, GT, FV, LP, CC, IGG) independently perused the description of treatments arms in the Methods section and isolated those with psychological components. Researchers noted whether the intervention appeared manualized and extracted any reference to a protocol or manual. Disagreements were resolved by consensus discussion with the senior author (IAC).

Supplementary material was also checked for any protocols included with the publication. For each reference, we examined if they contained a detailed description of the intervention. Second, trial registration numbers, when available, were extracted from each paper. Trial registrations were checked for the presence of any protocol. Third, for the studies where no detailed intervention descriptions were identified with the first two approaches, we extracted the names of the first, last and corresponding author. We queried Google for their current e-mail addresses. If no valid addresses were found, we searched for others from the author team. A common mail template (S1 Text) was developed, which included information about the objectives of the project (i.e., identifying active ingredients of psychological treatments) and a complete trial reference, and asked authors if they could provide the treatment protocols for the interventions used in the trial. The text explicitly stated that both published and unpublished protocols, such as the ones developed *ad hoc* for use in the trial, were valid sources of information. Emails were sent to first authors, followed by last and corresponding, depending on the e-mail addressed identified. In case of no reply, reminders we sent between 2 and 8 weeks after the first mail. We sent additional reminders if authors claimed to send materials subsequently and did not follow-back. If no reply was received, other authors were contacted. We also reached out to other authors, when those contacted indicated others had retained the protocol or manual. Fourth, if no treatment protocols were identified in steps 1-3, we searched Google for the manuals cited in the trial reports for each active intervention arm. For each protocol or manual retrieved, we checked if it was commercially or publicly available (i.e., defined as access with no restrictions, such as open-access manuals or publications, preprints, author version of records and others).

### Data extraction and synthesis

For each trial, we noted how many protocols or manuals were retrieved as (1) published (as supplement to the trial report or as separate publications), (2) from trial registries and (3) through author contact. These results were summarized by trial and by disorder, considering the protocol retrieved if it was available for at least one of the intervention arms. As some of the trials tested more than one psychological intervention, we also tabulated protocol or manual availability by intervention arms. We summed all arms with psychological components for each disorder and separately tabulated those not manualized. For step 4, we only report results by arm, given that in multi-arm trials, only some of the protocols would have to be retrieved by purchasing them, where others had already been obtained for example from author contact.

For each disorder, we computed the total number of active psychological intervention arms that were delivered according to protocols or manuals (1) *accessible*, i.e. retrieval from any source (published paper, trial registry, author contact, commercially available) and (2) *publicly available*.

Results are presented descriptively, as counts and proportions, with 95% confidence intervals (CI), calculated with the Clopper-Pearson method in Stata/SE version 16.1.

## Findings

### Selection of trials

From the 6 selected network meta-analyses, we identified 260 independent RCTs with a total of 422 psychological intervention arms. Figure 1 presents a PRISMA-type flow diagram of the selection and identification of the studies and protocols.

**Fig 1.**
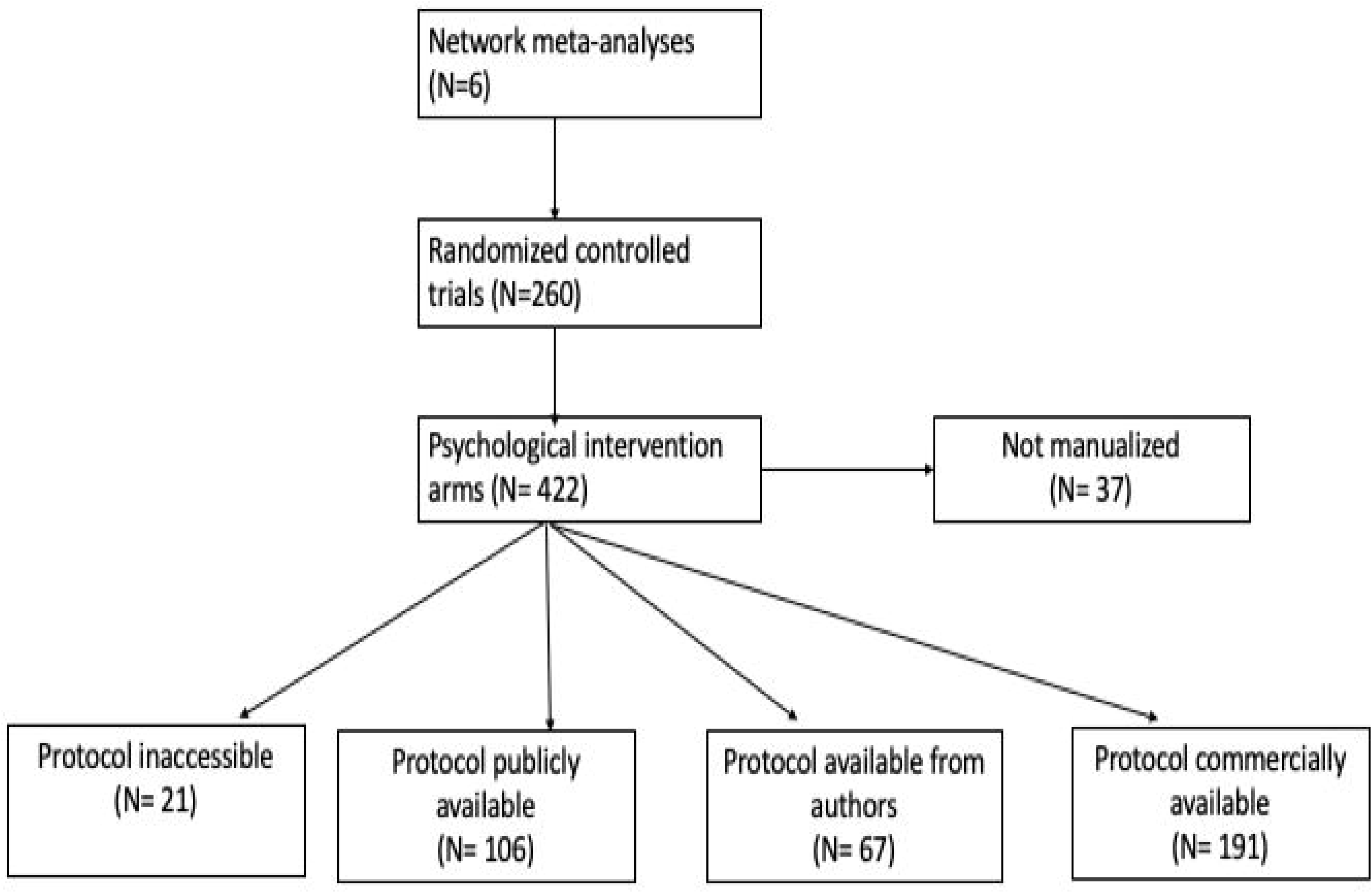
Flow diagram of study selection and retrieval of protocols and manuals for psychological intervention arms

### Retrieval of treatment protocols and manuals by trial

Steps 1 and 2 were conducted between January and April 2024. As shown in Table 1, we retrieved protocols (published independently or as supplements) for 20/260 trials (8%, 95% CI 5% to 20%). We identified trial registration numbers for 72/260 trials (28%, 95% CI 22% to 34%) (Table 1, S2 Table). For 12/260 studies (5%, 95% CI 2% to 8%), trial registries contained intervention protocols, all of which had been identified previously, leading to 20 trials (8%, 95% CI 5% to 12%) with intervention protocols after step 1 and 2 of our procedure, leading to 240 trials without published protocols for step 3.

**Table 1.** Retrieval of intervention descriptions from published protocols and trial registrations.

|  | **RCTs** | **Protocol**<br>**(published)<sup>i)</sup>** | **% protocol**<br>**published**<br>**(95% CI)<sup>ii)</sup>** | **Trial**<br>**registration** | **% registered**<br>**(95% CI)<sup>ii)</sup>** | **Description**<br>**(registration)** |
| --- | --- | --- | --- | --- | --- | --- |
| **Psychosis** | 93 | 5 | 5% (2% to 12%) | 21 | 23% (15% to 32%) | 3 |
| **BPD** | 43 | 5 | 12% (4% to 25%) | 16 | 37% (23% to 53%) | 3 |
| **SUD** | 50 | 1 | 2% (0.5% to 11%) | 13 | 26% (15% to 40%) | 0 |
| **Bipolar** | 40 | 6 | 15% (6% to 30%) | 11 | 27% (15% to 44%) | 4 |
| **Anorexia** | 13 | 3 | 23% (5% to 54%) | 6 | 46% (19% to 75%) | 2 |
| **Bulimia** | 21 | 0 | 0% | 5 | 24% (8% to 47%) | 0 |
| **TOTAL** | **260** | **20** | **8% (5% to 12%)** | **72** | **28% (22% to 34%)** | **12** |

*Note.* BPD, Borderline personality disorder; CI, confidence interval; SUD, Substance Use Disorder; RCT, randomized controlled trial
<sup>i)</sup> As supplement or as separate paper
<sup>ii)</sup> Out of total number of trials

In step 3, we could not identify any e-mail address for 11/240 trials (4%, 95% CI 2% to 8%). For the remaining 230 trials (229 trials without a protocol and one that was initially erroneously classified as having no published protocol), we contacted a total of 450 authors between April and July 2024. Author replies are classified in Table 2 and S3 Table.

**Table 2.**
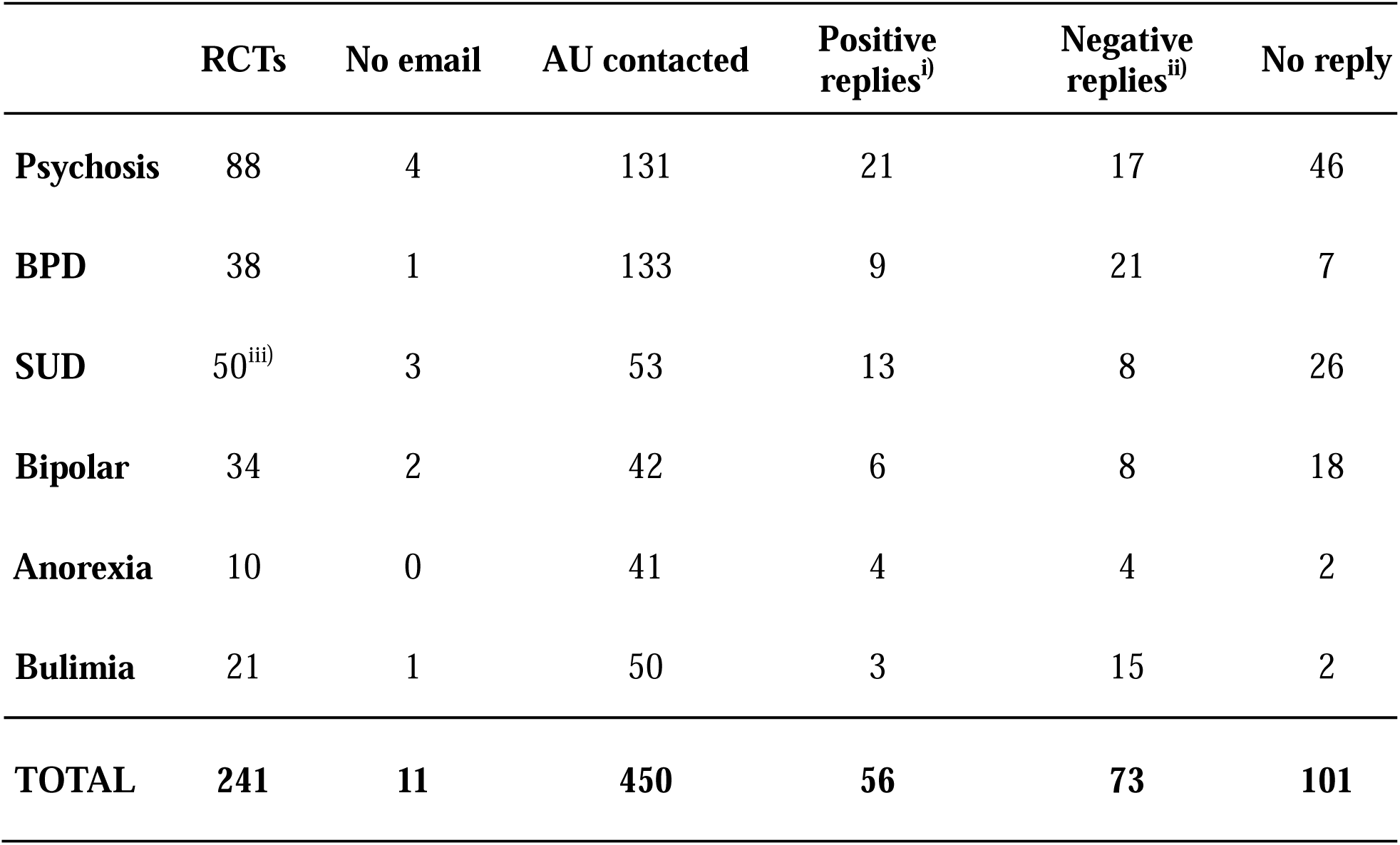
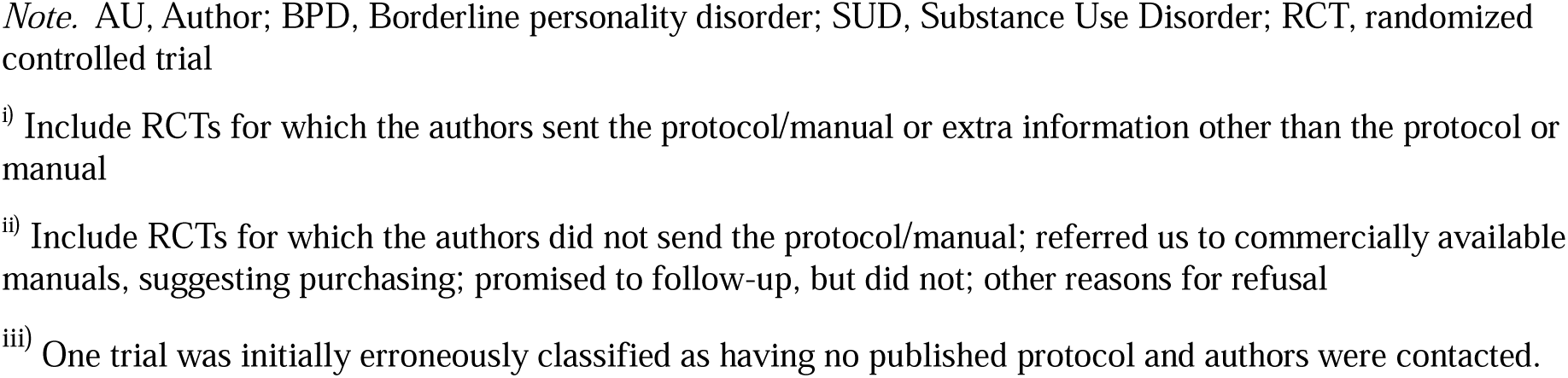
Retrieval of intervention descriptions through author contact.

|  | **RCTs** | **No email** | **AU contacted** | **Positive replies<sup>i)</sup>** | **Negative replies<sup>ii)</sup>** | **No reply** |
| --- | --- | --- | --- | --- | --- | --- |
| **Psychosis** | 88 | 4 | 131 | 21 | 17 | 46 |
| **BPD** | 38 | 1 | 133 | 9 | 21 | 7 |
| **SUD** | 50<sup>iii)</sup> | 3 | 53 | 13 | 8 | 26 |
| **Bipolar** | 34 | 2 | 42 | 6 | 8 | 18 |
| **Anorexia** | 10 | 0 | 41 | 4 | 4 | 2 |
| **Bulimia** | 21 | 1 | 50 | 3 | 15 | 2 |
| **TOTAL** | **241** | **11** | **450** | **56** | **73** | **101** |

*Note.* AU, Author; BPD, Borderline personality disorder; SUD, Substance Use Disorder; RCT, randomized controlled trial
i) Include RCTs for which the authors sent the protocol/manual or extra information other than the protocol or manual
ii) Include RCTs for which the authors did not send the protocol/manual; referred us to commercially available manuals, suggesting purchasing; promised to follow-up, but did not; other reasons for refusal
iii) One trial was initially erroneously classified as having no published protocol and authors were contacted.

We classified responses as positive for 56 of the 240 trials with no protocols (23%, 95% CI 18% to 29%). Of these, for 43/240 (18%, 95% CI 13% to 23%) trials, authors provided us with the protocol (in English or another language) or the manual of the intervention. For 13 trials (5%, 95% CI 3% to 9%), the authors sent additional information that did not contain the intervention protocol, manual or detailed description: extra references and papers (9), a PhD thesis (1), a Master thesis (1), a protocol from a trial registry (1), the document submitted for ethical approval (1)

We classified responses as negative for 73/240 trials (30%, 95% CI 25% to 37%). Of these, for 33 trials (14%, 95% CI 10% to 19%), the authors responded that the protocol was not available or that they had no access. For 21 trials (9%, 95% CI 5% to 13%), authors referred us to published, commercially available versions of the protocol or manual, suggesting we purchase these. For 13 trials (5%, 95% CI 3% to 9%), authors promised to follow-up, but never did, despite reminders, or forwarded our request to other members of the research team who would send the protocol. Finally, for 6 trials (3%, 95% CI 0.9% to 6%), authors refused to provide any materials for the following reasons: unspecified personal reasons (1), not involved in treatment development (1), intervention not manualized (1) protocol is retained by first author who cannot be contacted due to unexpected incident (1), no justification given (2). For the remaining 101 trials (42%, 95% CI 36% to 49%) we received no answer from any of the contacted authors, despite reminders.

Finally, in Step 4, we looked for commercially available manuals for the trials where we had identified nothing in previous steps. Public and commercial availability of manuals or protocols were checked between November 2024 and February 2025.

### Accessibility and public availability of treatment protocols by arm

As shown in S3 Table and Figure 1, 37 of 422 intervention arms (9%, 95% CI 6% to 12%) were explicitly non-manualized. Protocols or manuals were retrievable for 364/422 psychological intervention arms (86%, 95% CI 83% to 89%), considering all the approaches (published paper, trial registry, author contact, available commercially).

Of these, for 106/422 (25%, 95% CI 21% to 30%), protocols or manuals were publicly available and for 67/422 (16%, 95% CI 13% to 20%) we had obtained them from the authors. For 191 psychological intervention arms (45%, 95% CI 40% to 50%), protocols or manuals were only available commercially.

## Conclusions and clinical implications

In a large-scale evaluation of 422 evidence-based psychological interventions for severe mental disorders, studied in 260 randomized trials, sourced from 6 large network meta-analyses, we could retrieve a detailed intervention description, operationalized as a protocol or manual, for about 86% of the active interventions. Considering publicly available protocols or manuals only, this rate was reduced by more than 3 times, at 25%. Around 45% the protocols or manuals were only available commercially (as pay-walled publications or books that could be purchased). The retrieval rate is based on a resource-intensive, multipronged approach, which involved searching multiple sources (papers, trials registries, Google for commercially available manuals), as well as extensive author contact. Only 8% of the trials were associated to a protocol (published separately or as supplement), describing the intervention in detail. Under 30% of the trials were registered, and in only 5% of the trials the registration contained detailed intervention descriptions. Author contact led to the retrieval of an additional 43 protocols, amounting to around 20% of the trials with no protocol previously identified from reports and trial registries and 15% of the total psychological intervention arms. Of the 450 authors contacted, we received no reply, despite reminders, for around 40% of the trials. For another 30% of trials, authors declined to provide protocols or manuals, frequently suggesting we purchase them, though we had indicated our use was exclusively for research. For 5% of the trials, we could not identify contact data. Therefore, author contact led to positive, helpful replies for around a quarter of the trials.

We selected interventions studied in randomized controlled trials included in network meta-analyses. These allow for the combination of large collections of trials, even for interventions studied infrequently, and are considered the highest level of evidence in treatment guidelines^12^. Consequently, we can be reasonably certain that our analysis covered most evidence-based interventions for these disorders. Moreover, we considered severe mental disorders, for which there are often multiple bespoke interventions, based on distinct theoretical models and tested in few trials. Some of these interventions have specific and distinct components, not found in other treatments packages.

Like drugs, psychological interventions are used to treat mental disorders. Protocols and manuals could be viewed as analogous to the package inserts or labels in drug regulation, where the ingredients, indications, usage, dose, administration, and other details are specified. Just under 15% of the psychological intervention arms were either non-manualized or the manual was inaccessible (unpublished or otherwise unretrievable). For these, identifying active ingredients or reproducing the interventions are not possible. Our retrieval rate of 86% of protocols and manuals was time and resource-intensive, entailing an almost yearlong effort from a team of 7 researchers. A quarter of all psychological intervention arms had publicly available protocols or manuals. Contacting authors of trials testing the interventions, the most accessible retrieval method after public availability, produced modest results. Only 43 additional protocols were sent by authors, amounting to only 16% of all active intervention arms. Most authors were either not available or not willing to share, though it is possible they would have been more forthcoming in sharing protocols for clinical purposes. Therefore, reliance on open-access resources and sharing from authors would enable access to about 40% of trial protocols or manuals.

These findings are in stark contrast with recent calls for making manuals for psychological interventions freely available, particularly as many interventions were developed with public funds^21^. Public access to treatment manuals would greatly aid dissemination, particularly in low resourced settings, where access to and uptake of psychological treatments are woefully insufficient. For example, for schizophrenia there has been limited implementation of psychosocial interventions, with a median treatment gap of 69%, reaching 89% in low-income countries^22^. Conversely, commercialization of psychotherapy manuals through publishers could be seen as ensuring better distribution, thus aiding implementation and use. Estimating the cost of psychotherapy protocols and manuals, which would involve retrieving prices from various publishers and aggregate distributors like Amazon®, and comparing them to those of other treatments, like medication, was beyond the scope of the current work. These estimates would also not consider the cost of specialized training associated with many of the treatment manuals. However, regardless of the exact amount, the average cost will likely still be too high for low- and middle-income countries, where most people with mental disorders are located^21^. It was recently argued that treatment manuals for effective psychological treatments should be free or affordable, similarly to the drugs include in the World Health Organization Essential Medicines List (WHO-EML). It is unclear what proportion of all available drugs are on the WHO-EML, with one over 30-year old analysis estimating 16%^23^. Therefore, it is difficult to benchmark estimates of freely accessible psychological interventions against those for drugs. Still, for some disorders very few protocols or manuals were publicly available, e.g. 1 for bulimia, 5 for anorexia and 8 for borderline.

Without access to protocols or manuals, at least for explicit research purposes such as identifying active ingredients in the DECOMPOSE project, the content of psychological interventions is limited to descriptions in trial reports. If these descriptions are not complete or described at similar levels of detail, some components might be inadvertently merged (e.g., thought monitoring and cognitive restructuring) or missed entirely. This would severely constrain the potential of meta-analyses examining the differential efficacy of treatment components (“component network meta-analyses”^24^). Completeness of reporting for psychological interventions, for example as assessed with the Template for Intervention Description and Replication (TIDieR)^25^, received very little study. However, the few existent analyses showed inadequate reporting for interventions (including psychological) for alcohol use disorders^26^ and for psychological interventions for pain after knee replacement^27^. We could not identify any analysis of completeness of intervention reporting for other mental disorders. Similarly, examinations of the availability of treatment manuals for psychological interventions have been quasi non-existent, except for a small analysis of 27 trials set in low- and middle-income countries^10^. Of the 19 trials that reported using a manual, only 8 were referenced in the bibliography and only 2 were publicly available. However, the authors did not also attempt to retrieve manuals, as we have done here. One planned scoping review^28^ aims to characterize book-based psychotherapy manuals published up to 2022, with no results reported yet.

Our study has several limitations. First, we only focused on severe mental disorders, for which there are often many distinct psychological interventions, tested in few trials. Therefore, it is more likely that developers would be protective of the manuals and that there are fewer similar interventions, with overlapping components. Similar evaluations could be conducted for more common mental disorders, like depression or anxiety, where protocols are tested in more trials and where distinct protocols often share multiple components. Second, we did not contact all authors involved in the trial. However, given our extensive procedure, it is very likely we reached the lead authors or treatment developers. Third, we did not organize results by unique psychological interventions arms, as many trials described changes and adaptations of existent manuals, often making it challenging to ascertain if the treatment components remained unchanged. However, when the same intervention was used in more trials and the same manual was referenced, we considered it accessible if we had already retrieved for another trial. Fourth, we did not check whether other entities, such as funders, could have retained and publicly shared a copy of the intervention protocol.

Overall, our findings underscore the challenges of accessing detailed descriptions of psychological interventions, beyond what is reported in the paper, which in term hamper examinations of treatment components. Developing psychological treatments and evaluating these in RCTs need to be complemented by a more streamlined and less taxing access to treatment components and other characteristics (e.g., delivery modes, training required), similarly to labels of approved drugs. Furthermore, improving dissemination of effective psychological treatments, acknowledged as a global mental health priority^29^, requires public availability of a significant proportion of treatment manuals. For research purposes such as the DECOMPOSE project, funders could consider mandating sharing of treatment protocols. Some medical journals such as *Lancet* and *JAMA* family journal*, New England Journal of Medicine*, *Plos Medicine* or *Nature Medicine* currently mandate the inclusion of trial protocols for published randomized trials, a policy that should be generalized across journals. Trial registries could also require the inclusion of the trial protocol. However, it would be important to make sure intervention descriptions in trial protocols are detailed and complete. More systematic investigations into completeness of reporting of psychological interventions for mental disorders, for example using TIDieR, could provide an estimate of the feasibility of identifying active ingredients based on what is reporting in the paper or in trial protocols shared with publication. Finally, systematic investigation into the accessibility of treatment manuals and protocols for other disorders are necessary to establish the proportion of evidence-based psychological interventions described with sufficient detail to enable identification of active ingredients.

## Supporting information

PRISMA Checklist

## Data Availability

All metadata and data extracted for this study, except for personal data (name and e-mail contact of authors contacted), along with the data dictionary, are publicly available on the Zenodo repository DOI: 10.5281/zenodo.14543415.

https://doi.org/10.5281/zenodo.14543415

## Ethics approval and consent to participate

Not applicable

## Consent for publication

Not applicable

## Authors’ contributions

Conceptualization: IAC

Data curation: CB, GT, FV, LP, CC, IGG

Formal analysis: CB, IAC

Funding acquisition: IAC

Investigation: CB, GT, FV, LP, CC, IGG

Methodology: IAC

Project administration: IAC

Supervision: IAC, CB, GT

Validation: CB, GT, FV, LP, CC, IGG

Visualization: CB, IGG

Writing-original draft: IAC, CB

Writing-review and editing: GT, FV, LP, CC, IGG

IAC is the guarantor for the study.

