## Supplementary material for "Access to treatment protocols and manuals for evidence-based psychological interventions for severe mental disorders: A survey of randomized trials included in network meta-analyses"

**Appendix**

**S1 Text. E-mail Template for protocol retrieval**

Dear Prof. [Insert name]

We are a team of researchers working on the project ***Disentangling psychological interventions for mental disorders into a taxonomy of active ingredients (DECOMPOSE)****,* principal investigator Prof. Ioana Cristea, University of Padova, Italy.

The project is funded by an European Research Council (ERC) Starting Grant under the grant agreement 101042701.

In this project, we aim to dismantle psychological interventions for several mental disorders into their constituent parts, integrate components into a taxonomy, and recast treatment efficacy and personalization from a novel perspective: components instead of brands and categories. The final goal is to create an open clinical decision support system, where users can “assemble” and “dismantle” interventions, visualising gain or loss of treatment effectiveness.

Please see [here](https://cordis.europa.eu/project/id/101042701) and [here](https://www.bps.org.uk/psychologist/what-inside-treatment) for more details

In order to identify active ingredients, we are looking for **trial protocols that describe the intervention used in the trial**. It does not matter if these protocols were published or simply used for internal communication purposes in the context of the trial, because we only use protocols so as to have a detailed description of what was done during the intervention. From these descriptions, various rounds of data extraction, coding, and Delphi panels will work to identify treatment components.

Once components have been extracted, we will share the list with you and consult you as to whether you find the list accurate or there are omissions.

We have selected your trial [Full Reference] for inclusion.

Would you kindly share with us the protocol for the following treatment arms [insert complete non-abbreviated name of treatment]? This protocol can be in any format you have it available.

Thank you for your help!

[Name and Degree, On Behalf of the DECOMPOSE team]

**S1 Table. Identification and selection of included network meta-analyses**

| Disorder | Search string | Hits | Eligible NMAs | Reasons for rejection |
| --- | --- | --- | --- | --- |
| Psychosis | "network meta analysis"[Title/Abstract] AND ("psychotic"[Title/Abstract] OR "schizophrenia"[Title/Abstract]) AND ("psychol*"[Title/Abstract] OR "psychosocial*"[Title/Abstract] OR "psychother*"[Title/Abstract]) AND 2018/11/01:2023/11/01[Date - Publication] | 10 | 5 | 1 on relapse prevention  1 on stigma  1 children and adolescents  1 on a specific type of intervention |
| Borderline personality disorder | ((network meta-analysis[Title/Abstract]) AND (borderline[Title/Abstract]) AND (psychol*[Title/Abstract] OR psychosocial*[Title/Abstract] OR psychother*[Title/Abstract])) AND (("2018/11/1"[Date - Publication] : "2023/11/1"[Date - Publication])) | 2 | 1 | Only one eligible hit |
| Substance-used disorder | "network meta analysis"[Title/Abstract] AND ("alcohol"[Title/Abstract] OR "opioid"[Title/Abstract] OR "cannabis"[Title/Abstract] OR "hallucinogens"[Title/Abstract] OR "inhalants"[Title/Abstract] OR "sedatives"[Title/Abstract] OR "hypnotics"[Title/Abstract] OR "anxiolytics"[Title/Abstract] OR "stimulants"[Title/Abstract] OR "amphetamine"[Title/Abstract] OR "cocaine"[Title/Abstract] OR "substance"[Title/Abstract]) AND ("psychol*"[Title/Abstract] OR "psychosocial*"[Title/Abstract] OR "psychother*"[Title/Abstract]) AND 2018/11/01:2023/11/01[Date - Publication] ^1^ | 14 | 6 | 1 harmful alcohol use, not alcohol use disorder  1 on opioid-dependent patients on methadone maintenance  1 smaller number of included trials than selected NMA  1 comorbid alcohol use and depression  1 included interventions also pharmacological for maintaining abstinence |
| Bipolar disorder | "network meta analysis"[Title/Abstract] AND ("bipolar"[Title/Abstract] OR "mood"[Title/Abstract]) AND ("psychol*"[Title/Abstract] OR "psychosocial*"[Title/Abstract] OR "psychother*"[Title/Abstract]) AND 2018/11/01:2023/11/01 [Date - Publication] | 16 | 2 | 1 on a specific type of intervention for depressive symptoms |
| Anorexia nervosa/bulimia nervosa | ((network meta-analysis[Title/Abstract]) AND (anorexia[Title/Abstract] OR bulimia[Title/Abstract] OR eating[Title/Abstract])) AND (("2018/11/01"[Date - Publication] : "2023/11/01"[Date - Publication]))^2^ | 2 | 2 | 1 eligible hit for each disorder |

*Note.*

^1^ Caffeine and tobacco use were excluded

^2^ Terms related to the type of interventions were eliminated from the search string because no hits were initially retrieved.

**S2 Table. Available information in trial registries**

| Disorder | Number | Trial reference | Trial registration number | Info retrieved from registration |
| --- | --- | --- | --- | --- |
| Psychosis  N=21 | 1 | Chien WT, Thompson DR. Effects of a mindfulness-based psychoeducation programme for Chinese patients with schizophrenia: 2-year follow-up. Br J Psychiatry. 2014;205:52–9. | NCT01667601 | No detailed description |
|  | 2 | Dickinson D, Ph D, Tenhula W, Ph D, Morris S, Ph D, et al. A randomized , controlled trial of computer-assisted cognitive remediation for schizophrenia. Am J Psychiatry. 2010;167:170–80. | NCT00295048, NCT00261794 | No detailed description |
|  | 3 | Farreny A, Aguado J, Ochoa S, Huerta-ramos E, Marsà F, López-carrilero R, et al. REPYFLEC cognitive remediation group training in schizophrenia Looking for an integrative approach. Schizophr Res [Internet]. Elsevier B.V.; 2012;142(1–3):137–44. Available from: http://dx.doi.org/10.1016/j.schres.2012.08.036 | NCT01279070 | No detailed description |
|  | 4 | Freeman 2015a: Freeman D, Dunn G, Startup H, Pugh K, Cordwell J, Mander H, et al. Effects of cognitive behaviour therapy for worry on persecutory delusions in patients with psychosis (WIT): a parallel, single-blind , randomised controlled trial with a mediation analysis. Lancet Psychiatry. 2015;2(April 2015):305–13. | ISRCTN23197625 | Protocol published separately |
|  | 5 | Freeman 2015b: Freeman D, Waite F, Startup H, Myers E, Lister R, Mcinerney J, et al. Efficacy of cognitive behavioural therapy for sleep improvement in patients with persistent delusions and hallucinations (BEST): a prospective , assessor-blind , randomised controlled pilot trial. Lancet Psychiatry. 2015;2:975–83. | ISRCTN 33695128 | Protocol published separately |
|  | 6 | Garety 2008 (i) & (ii): Garety PA, Fowler DG, Freeman D, Bebbington P, Dunn G, Kuipers E. Cognitive – behavioural therapy and family intervention for relapse prevention and symptom reduction in psychosis : randomised controlled trial. Br J Psychiatry. 2008;192:412–23. | ISRCTN83557988 | No detailed description |
|  | 7 | Guo X, Zhai J, Liu Z, Fang M, Wang B, Wang C, et al. Antipsychotic medication alone versus combined with psychosocial intervention on outcomes of early stage schizophrenia: a randomized, one-year study. Arch Gen Psychiatry. 2010;67(9):895–904. | NCT00654576 | No detailed description |
|  | 8 | Jørgensen R, Licht RW, Lysaker PH, Munk-Jørgensen P, Buck KD, Jensen SOW, et al. Effects on cognitive and clinical insight with the use of Guided Self-Determination in outpatients with schizophrenia : A randomized open trial. Eur Psychiatry. 2015;30:655–63. | NCT01282307 | No detailed description |
|  | 9 | Kantrowitz JT, Sharif Z, Medalia A, Keefe RSE, Harvey P, Bruder G, et al. A multicenter, rater-blinded, randomised controlled study of auditory processing-focused cognitive remediation combined with open-label Lurasidone in patients with schizphrenia and schizoaffective disorder. J Clin Psychiatry. 2016;77(6):799–806. | NCT01173874 | No detailed description |
|  | 10 | Keefe RSE, Vinogradov S, Medalia A, Peter F, Caroff SN, Souza DCD, et al. Feasibility and pilot efficacy results from the multi-site Cognitive Remediation in the Schizophrenia Trials Network (CRSTN) Study. J Clin Psychiatry. 2012;73(7):1016–22. | NCT00930150 | No detailed description |
|  | 11 | Li Z, Guo Z, Wang N, Xu Z, Qu Y, Wang X, et al. Cognitive – behavioural therapy for patients with schizophrenia : a multicentre randomized controlled trial in Beijing , China. Psychol Med. 2015;45:1893–905. | ChiCTR-TRC-08000124 | No detailed description |
|  | 12 | Lincoln TM, Ziegler M, Mehl S, Kesting M, Lullmann E, Westermann S, et al. Moving from efficacy to effectiveness in Cognitive Behavioral Therapy for Psychosis : A randomized clinical practice trial. J Consult Clin Psychol. 2012;80(4):674–86. | NCT00300651 | No detailed description |
|  | 13 | Moritz S, Veckenstedt R, Randjbar S, Vitzthum F, Woodward TS. Antipsychotic treatment beyond antipsychotics: metacognitive intervention for schizophrenia patients improves delusional symptoms. Psychol Med. 2011;41:1823–32. | NCT01029067 | No detailed description |
|  | 14 | Moritz S, Veckenstedt R, Bohn F, Hottenrott B, Scheu F, Randjbar S, et al. Complementary group Metacognitive Training (MCT) reduces delusional ideation in schizophrenia. Schizophr Res [Internet]. Elsevier B.V.; 2013;151(1–3):61–9. Available from: http://dx.doi.org/10.1016/j.schres.2013.10.007 | ISRCTN95205723 | No detailed description |
|  | 15 | Morrison AP, Turkington D, Pyle M, Spencer H, Brabban A, Dunn G, et al. Cognitive therapy for people with schizophrenia spectrum disorders not taking antipsychotic drugs : a single-blind randomised controlled trial. Lancet [Internet]. Elsevier Ltd; 2014: 383(9926):1395–403. Available from: http://dx.doi.org/10.1016/S0140-6736(13)62246-1 | ISRCTN29607432 | No detailed description |
|  | 16 | Naeem F, Saeed S, Irfan M, Kiran T, Mehmood N, Gul M, et al. Brief culturally adapted CBT for psychosis (CaCBTp): A randomized controlled trial from a low income country. Schizophr Res. 2015;164:143–8. | NCT01876056 | No detailed description |
|  | 17 | Rathod S, Phiri P, Harris S, Underwood C, Thagadur M, Padmanabi U, et al. Cognitive behaviour therapy for psychosis can be adapted for minority ethnic groups : A randomised controlled trial. Schizophr Res [Internet]. Elsevier B.V.; 2013;143(2–3):319–26. Available from: http://dx.doi.org/10.1016/j.schres.2012.11.007 | ISRCTN95603741 | No detailed description |
|  | 18 | Schrank B, Brownell T, Jakaite Z, Larkin C, Pesola F, Riches S, Tylee A, Slade M. Evaluation of a positive psychotherapy group intervention for people with psychosis: pilot randomised controlled trial. Epid Psychiatric Sci. 2016; 25: 235–246. | ISRCTN04199273 | Protocol published separately |
|  | 19 | Velligan DI, Tai S, Roberts DL, Maples-Aguilar N, Brown M, Mintz J, et al. A randomized controlled trial comparing cognitive behavior therapy , cognitive adaptation training , their combination and treatment as usual in chronic schizophrenia. Schizophr Bull. 2015;41(3):597–603. | NCT01915017) | No detailed description |
|  | 20 | Wang L-Q, Chien WT, Yip LK, Karatzias T. A randomized controlled trial of a mindfulness- based intervention program for people with schizophrenia : 6-month follow-up. Neuropsychiatr Dis Treat. 2016;12:3097–110. | NCT01667601 | No detailed description |
|  | 21 | Wykes T, Reeder C, Landau S, Everitt B, Knapp M, Patel A, Romeo R. Cognitive remediation therapy in schizophrenia. Br J Psychiatry, 2007; 190, 421–427. | ISRCTN44277627 | No detailed description |
| Borderline Personality Disorder  N=16 | 1 | Amianto, F., Ferrero, A., Pierò, A. et al. Supervised team management, with or without structured psychotherapy, in heavy users of a mental health service with borderline personality disorder: a two-year follow-up preliminary randomized study. BMC Psychiatry 11, 181 (2011). https://doi.org/10.1186/1471-244X-11-181 | NCT1356069 | No detailed description |
|  | 2 | Andreasson K, Krogh J, Wenneberg C, Jessen HK, Krakauer K, Gluud C, Thomsen RR, Randers L, Nordentoft M. Effectiveness of dialectical behavior therapy versus collaborative assessment and management of suicidality treatment for reduction of self-harm in adults with borderline personality traits and disorder-a randomized observer-blinded clinical triaL. Depress Anxiety. 2016 Jun;33(6):520-30. doi: 10.1002/da.22472. | NCT01512602 | Protocol published separately |
|  | 3 | Bateman A, Fonagy P. Randomized controlled trial of outpatient mentalization-based treatment versus structured clinical management for borderline personality disorder. Am J Psychiatry. 2009 Dec;166(12):1355-64. doi: 10.1176/appi.ajp.2009.09040539. | ISRCTN27660668 | No detailed description |
|  | 4 | Bohus M, Kleindienst N, Hahn C, Müller-Engelmann M, Ludäscher P, Steil R, Fydrich T, Kuehner C, Resick PA, Stiglmayr C, Schmahl C, Priebe K. Dialectical Behavior Therapy for Posttraumatic Stress Disorder (DBT-PTSD) Compared With Cognitive Processing Therapy (CPT) in Complex Presentations of PTSD in Women Survivors of Childhood Abuse: A Randomized Clinical Trial. JAMA Psychiatry. 2020 Dec 1;77(12):1235-1245. doi: 10.1001/jamapsychiatry.2020.2148. | DRKS00005578 | No detailed description |
|  | 5 | Bozzatello, P., & Bellino, S. (2020). Interpersonal psychotherapy as a single treatment for borderline personality disorder: A pilot randomized-controlled study. Frontiers in Psychiatry, 11, 578910. doi:10.3389/fpsyt.2020.578910 | ACTRN12619000078156 | No detailed description |
|  | 6 | Carlyle, D., Green, R., Inder, M., Porter, R., Crowe, M., Mulder, R., & Frampton, C. (2020). A randomized-controlled trial of mentalization-based treatment compared with structured case management for borderline personality disorder in a mainstream public health service. Frontiers in Psychiatry, 11, 561916. doi:10.3389/fpsyt.2020.561916 | ACTRN12612000951853 | No detailed description |
|  | 7 | Crawford, M. J., Thana, L., Parker, J., Turner, O., Carney, A., McMurran, M., … Spong, A. (2020). Structured psychological support for people with personality disorder: Feasibility randomised controlled trial of a low-intensity intervention. British Journal of Psychiatry Open, 6(2), e25. doi:10.1192/ bjo.2020.7 | ISRCTN14994755 | Protocol published separately |
|  | 8 | Doering S, Hörz S, Rentrop M, Fischer-Kern M, Schuster P, Benecke C, Buchheim A, Martius P, Buchheim P. Transference-focused psychotherapy v. treatment by community psychotherapists for borderline personality disorder: randomised controlled trial. Br J Psychiatry. 2010 May;196(5):389-95. doi: 10.1192/bjp.bp.109.070177. | NCT00714311 | No detailed description |
|  | 9 | Herpertz, S. C., Matzke, B., Hillmann, K., Neukel, C., Mancke, F., Jaentsch, B., Schwenger, U., Honecker, H., Bullenkamp, R., Steinmann, S., Krauch, M., Bauer, S., Borzikowsky, C., Bertsch, K., & Dempfle, A. (2020). A mechanism-based group-psychotherapy approach to aggressive behaviour in borderline personality disorder: findings from a cluster-randomised controlled trial. BJPsych open, 7(1), e17. https://doi.org/10.1192/bjo.2020.131 | DRKS00009445 | Protocol included in trial report |
|  | 10 | Hilden, H., Rosenström, T., Karila, I., Elokorpi, A., Torpo, M., Arajärvi, R., & Isometsä, E. (2020). Effectiveness of brief schema group therapy for borderline personality disorder symptoms: a randomized pilot study. Nordic Journal of Psychiatry, 75(3), 176–185. https://doi.org/10.1080/08039488.2020.1826050 | ISRCTN76381242 | No detailed description |
|  | 11 | Laurenssen, E. M. P., Luyten, P., Kikkert, M. J., Westra, D., Peen, J., Soons, M. B. J., van Dam, A. M., van Broekhuyzen, A. J., Blankers, M., Busschbach, J. J. V., & Dekker, J. J. M. (2018). Day hospital mentalization-based treatment v. specialist treatment as usual in patients with borderline personality disorder: randomized controlled trial. Psychological medicine, 48(15), 2522–2529. https://doi.org/10.1017/S0033291718000132 | NTR2175 | No detailed description |
|  | 12 | McMain, S. F., Links, P. S., Gnam, W. H., Guimond, T., Cardish, R. J., Korman, L., & Streiner, D. L. (2009). A randomized trial of dialectical behavior therapy versus general psychiatric management for borderline personality disorder. The American journal of psychiatry, 166(12), 1365–1374. https://doi.org/10.1176/appi.ajp.2009.09010039 | NCT00154154 | No detailed description |
|  | 13 | McMain, S. F., Guimond, T., Barnhart, R., Habinski, L., & Streiner, D. L. (2017). A randomized trial of brief dialectical behaviour therapy skills training in suicidal patients suffering from borderline disorder. Acta psychiatrica Scandinavica, 135(2), 138–148. https://doi.org/10.1111/acps.12664 | NCT01193205 | No detailed description |
|  | 14 | Philips, B., Wennberg, P., Konradsson, P., & Franck, J. (2018). Mentalization-Based Treatment for Concurrent Borderline Personality Disorder and Substance Use Disorder: A Randomized Controlled Feasibility Study. European addiction research, 24(1), 1–8. https://doi.org/10.1159/000485564 | ISRCTN98982683 | No detailed description |
|  | 15 | Stanley, B. (2017). Comparing treatments for self-injury and suicidal behavior in people with borderline personality disorder [published data only]. NCT00834834. | NCT00834834 | No detailed description |
|  | 16 | Walton, C. J., Bendit, N., Baker, A. L., Carter, G. L., & Lewin, T. J. (2020). A randomised trial of dialectical behaviour therapy and the conversational model for the treatment of borderline personality disorder with recent suicidal and/or non-suicidal self-injury: An effectiveness study in an Australian public mental health service. The Australian and New Zealand journal of psychiatry, 54(10), 1020–1034. https://doi.org/10.1177/0004867420931164 | ACTRN12612001187831 | No detailed description |
| Substance Use Disorder n= 13 | 1 | Carroll KM, Nich C, Shi JM, Eagan D, Ball SA. Efficacy of disulfiram and Twelve Step Facilitation in cocaine-dependent individuals maintained on methadone: a randomized placebo-controlled trial. Drug Alcohol Depend 2012;126(1-2):224-231. doi: 10.1016/j.drugalcdep.2012.05.019 pmid: 22695473 | NCT00350870 | No detailed description |
|  | 2 | Carroll KM, Kiluk BD, Nich C, et al. Computer-assisted delivery of cognitive-behavioral therapy: efficacy and durability of CBT4CBT among cocaine-dependent individuals maintained on methadone. Am J Psychiatry 2014;171(4):436-444. doi: 10.1176/appi.ajp.2013.13070987 pmid: 24577287 | NCT00350610 | No detailed description |
|  | 3 | ·Carroll KM, Nich C, Petry NM, Eagan DA, Shi JM, Ball SA. A randomized factorial trial of disulfiram and contingency management to enhance cognitive behavioral therapy for cocaine dependence*. Drug Alcohol Depend* 2016;160:135-142. doi: 10.1016/j.drugalcdep.2015.12.036 pmid: 26817621 | NCT00350870 | No detailed description |
|  | 4 | Donovan DM, Daley DC, Brigham GS, et al. Stimulant abuser groups to engage in 12-step: a multisite trial in the National Institute on Drug Abuse Clinical Trials Network. J Subst Abuse Treat 2013;44(1):103-114. doi: 10.1016/j.jsat.2012.04.004 pmid: 22657748 | NCT00573183. | No detailed description |
|  | 5 | Landovitz RJ, Fletcher JB, Shoptaw S, Reback CJ. Contingency management facilitates the use of postexposure prophylaxis among stimulant-using men who have sex with men *Open Forum Infect Dis*. 2015;2(1):ofu114. doi: 10.1093/ofid/ofu114 pmid: 25884003 | NCT01140880 | No detailed description |
|  | 6 | McDonell MG, Srebnik D, Angelo F, et al. Randomized controlled trial of contingency management for stimulant use in community mental health patients with serious mental illness*. Am J Psychiatry* 2013;170(1):94-101. doi: 10.1176/appi.ajp.2012.11121831 pmid: 23138961 | NCT00809770 | No detailed description |
|  | 7 | Menza TW, Jameson DR, Hughes JP, Colfax GN, Shoptaw S, Golden MR. Contingency management to reduce methamphetamine use and sexual risk among men who have sex with men: a randomized controlled trial*. BMC Public Health* 2010;10:774-2458-10-774. doi: 10.1186/1471-2458-10-774 pmid: 21172026 | NCT01174654 | No detailed description |
|  | 8 | Miguel AQ, Madruga CS, Cogo-Moreira H, et al. Contingency management is effective in promoting abstinence and retention in treatment among crack cocaine users in Brazil: A randomized controlled trial. Psychol Addict Behav 2016;30(5):536-543. doi:10.1037/adb0000192 pmid: 27442691 | NCT01815645. | No detailed description |
|  | 9 | Milby JB, Schumacher JE, Vuchinich RE, Freedman MJ, Kertesz S, Wallace D. Toward cost-effective initial care for substance-abusing homeless. J Subst Abuse Treat 2008;34(2):180-191. doi: 10.1016/j.jsat.2007.03.003 pmid: 17512156 | NCT00368524 | No detailed description |
|  | 10 | Petitjean SA, Dursteler-MacFarland KM, Krokar MC, et al. A randomized, controlled trial of combined cognitive-behavioral therapy plus prize-based contingency management for cocaine dependence. Drug Alcohol Depend 2014;145:94-100. doi: 10.1016/j.drugalcdep.2014.09.785 pmid: 25456571 | NCT00877435 | No detailed description |
|  | 11 | Petry NM, Alessi SM, Rash CJ. A randomized study of contingency management in cocaine-dependent patients with severe and persistent mental health disorders*. Drug Alcohol Depend* 2013;130(1-3):234-237. doi:10.1016/j.drugalcdep.2012.10.017 pmid: 23182410 | NCT01478815 | No detailed description |
|  | 12 | Schottenfeld RS, Moore B, Pantalon MV. Contingency management with community reinforcement approach or twelve-step facilitation drug counseling for cocaine dependent pregnant women or women with young children. Drug Alcohol Depend 2011;118(1):48-55. doi: 10.1016/j.drugalcdep.2011.02.019 pmid: 21454024 | NCT00914381 | No detailed description |
|  | 13 | Umbricht A, DeFulio A, Winstanley EL, et al. Topiramate for cocaine dependence during methadone maintenance treatment: a randomized controlled trial. Drug Alcohol Depend 2014;140:92-100. doi: 0.1016/j.drugalcdep.2014.03.033 pmid: 24814607 | NCT00685178 | No detailed description |
| Bipolar Disorder  N=11 | 1 | Inder ML, Crowe MT, Luty SE, et al. Randomized, controlled trial of Interpersonal and Social Rhythm Therapy for young people with bipolar disorder. Bipolar Disord. 2015;17(2):128-138. | ACTRN12605000722695 | No detailed description |
|  | 2 | Jones SH, Smith G, Mulligan LD, et al. Recovery-focused cognitive-behavioural therapy for recent-onset bipolar disorder: randomised controlled pilot trial. Br J Psychiatry. 2015;206(1):58-66. | ISRCTN43062149 | No detailed description |
|  | 3 | Lobban F, Taylor L, Chandler C, et al. Enhanced relapse prevention for bipolar disorder by community mental health teams: cluster feasibility randomised trial. Br J Psychiatry. 2010;196(1):59-63. | ISRCTN41352631 | No detailed description |
|  | 4 | Miklowitz DJ, Otto MW, Frank E, et al. Psychosocial treatments for bipolar depression: a 1-year randomized trial from the Systematic Treatment Enhancement Program. Arch Gen Psychiatry. 2007;64:419-427. | NCT00012558 | No detailed description |
|  | 5 | Miklowitz DJ, Schneck CD, George EL, et al. Pharmacotherapy and family-focused treatment for adolescents with bipolar I and II disorders: a 2-Year randomized trial. Am J Psychiatry. 2014;171(6):658-667. | NCT00332098 | No detailed description |
|  | 6 | Morriss R, Lobban F, Riste L, et al. Clinical Effectiveness and Acceptability of Structured Group Psychoeducation Versus Optimised Unstructured Peer Support for Patients With Remitted Bipolar Disorder (PARADES): A Pragmatic, Multicentre, Observer-Blind, Randomised Controlled Superiority Trial. Lancet Psychiatry. 2016;3(11):1029-1038. | ISRCTN62761948 | Protocol published separately |
|  | 7 | Parikh SV, Zaretsky A, Beaulieu S, et al. A randomized controlled trial of psychoeducation or cognitive-behavioral therapy in bipolar disorder: a Canadian Network for Mood and Anxiety treatments (CANMAT) study. J Clin Psychiatry. 2012;73(6):803-810. | NCT00188838 | No detailed description |
|  | 8 | Perich T, Manicavasagar V, Mitchell PB, Ball JR, Hadzi-Pavlovic D. A randomized controlled trial of mindfulness-based cognitive therapy for bipolar disorder. Acta Psychiatr Scand. 2013;127(5):333-343. | ACTRN12610000414011 UTN 1111-1115-0862 | No detailed description |
|  | 9 | Torrent C, del Mar Bonnin C, Martinez-Aran A, et al. Efficacy of functional remediation in bipolar disorder: a multicenter randomized controlled study. Am J Psychiatry. 2013;170(8):852-859. | NCT01370668 | Protocol included with trial report |
|  | 10 | van der Voort TYG, van Meijel B, Goossens PJJ, et al. Collaborative care for patients with bipolar disorder: randomised controlled trial. Br J Psychiatry. 2015;206(5):393-400. | NTR2600 | Protocol included with trial report |
|  | 11 | Hautzinger M; A2 BipoLife Consortium. Adjuvant Psychotherapies to Prevent Relapse in Bipolar Disorder: A Randomized Clinical Trial. JAMA Psychiatry. 2024 Jun 5:e241310. doi: 10.1001/jamapsychiatry.2024.1310. Epub ahead of print. PMID: 38837133; PMCID: PMC11154370. | NCT02506322 | Protocol included with trial report |
| Anorexia  N=6 | 1 | Byrne S, Wade T, Hay P, et al. A randomised controlled trial of three psychological treatments for anorexia nervosa. Psychol Med 2017; 47: 2823–33 | ACTRN12611000725965 | No detailed description |
|  | 2 | Hay P, Touyz S, Arcelus J, et al. A randomized controlled trial of the compuLsive Exercise Activity TheraPy (LEAP): a new approach to compulsive exercise in anorexia nervosa. Int J Eat Disord 2018; 51: 999–1004 | ACTRN12610000585022 | Somewhat detailed description |
|  | 3 | Schmidt U, Magill N, Renwick B, et al. The Maudsley Outpatient Study of Treatments for Anorexia Nervosa and Related Conditions (MOSAIC): comparison of the Maudsley model of anorexia nervosa treatment for adults (MANTRA) with specialist supportive clinical management (SSCM) in outpatients with broadly defined anorexia nervosa: a randomized controlled trial. J Consult Clin Psychol 2015; 83: 796–807 | ISRCTN67720902 | Protocol published separately |
|  | 4 | Touyz S, Le Grange D, Lacey H, et al. Treating severe and enduring anorexia nervosa: a randomized controlled trial. Psychol Med 2013; 43: 2501–11 | ACTRN12607000440426 | No detailed description |
|  | 5 | Zipfel S, Wild B, Gross G, et al. Focal psychodynamic therapy, cognitive behaviour therapy, and optimised treatment as usual in outpatients with anorexia nervosa (ANTOP study): randomised controlled trial. Lancet 2014; 383: 127–37 | ISRCTN72809357 | Protocol published separately |
|  | 6 | Schmidt U, Oldershaw A, Jichi F, et al. Out-patient psychological therapies for adults with anorexia nervosa: randomised controlled trial. *British Journal of Psychiatry*. 2012;201(5):392-399. doi:10.1192/bjp.bp.112.112078 | ISRCTN62920529 | No detailed description. |
| Bulimia  N= 5 | 1 | Fairburn et al., 2009 - Transdiagnostic cognitive-behavioral therapy for patients with eating disorders: a two-site trial with 60-week follow-up | ISRCTN15562271 | No detailed description |
|  | 2 | Fairburn et al., 2015 - A transdiagnostic comparison of enhanced cognitive behaviour therapy (CBT-E) and interpersonal psychotherapy in the treatment of eating disorders. | ISRCTN15562271 | No detailed description |
|  | 3 | Sanchez-Ortiz et al., 2011 - A randomized controlled trial of internet-based cognitive-behavioural therapy for bulimia nervosa or related disorders in a student population | ISRCTN07388346 | No detailed description |
|  | 4 | Thompson‐Brenner et al., 2016 - Focused vs. Broad enhanced cognitive behavioral therapy for bulimia nervosa with comorbid borderline personality: A randomized controlled trial. | NCT00494858 | No detailed description |
|  | 5 | Wagner et al., 2013 - Internet-delivered cognitive–behavioural therapy v. conventional guided self-help for bulimia nervosa: long-term evaluation of a randomised controlled trial. | NCT00461071 | No detailed description |

**S3 Table. Classification of replies received from authors contacted**

|  | **Reply categories** | RCTs |
| --- | --- | --- |
| **Positive** | Sent the protocols or the manual | 43 |
|  | Additional information that did not contain the intervention protocol, manual or detailed description | 13 |
| **Negative** | No access to the protocol/data not available/paper is the only resource | 33 |
|  | They would follow up but never did /Forward email to other authors, who also never replies | 13 |
|  | Referred us to the published manuals /suggested purchasing them | 21 |
|  | Other | 6 |

**S4 Table.** Accessibility and public availability of treatment protocols and manuals by active intervention arms

|  | **RCTs** | **Active**  **arms^a^** | **Not manualized** | **Non manualized % (95% CI) ^c^** | **Inaccessible (unpublished)** | **Inaccessible %** | **Accessible^b^** | **Accessible %**  **(95% CI)^c^** | **Public^d^** | **Public %**  **(95% CI)^c^** | **Author^e^** | **Author ^e^**  **%**  **(95% CI)^c^** | **Commercial^f^** | **Commercial^f^**  **%**  **(95% CI)^c^** |
| --- | --- | --- | --- | --- | --- | --- | --- | --- | --- | --- | --- | --- | --- | --- |
| **Psychosis** | 93 | 123 | 10 | 8% (4% to 14%%) | 15 (5) | 12% (7% to 19%) | 98 | 80% (71% to 86%) | 27 | 22% (15% to 30%) | 14 | 11% (6% to 18%) | 57 | 46% (37% to 56%) |
| **BPD** | 43 | 62 | 2 | 3% (0.3% to 11%) | 2 (2) | 3% (0.4% to 11%) | 58 | 94% (84% to 98%) | 8 | 13% (6% to 24%) | 10 | 16% (8% to 28%) | 40 | 64% (51% to 76%) |
| **SUD^g^** | 50 | 104 | 9 | 9% (4% to 16%) | 0 | 0 | 95 | 91% (84% to 96%) | 44 | 42% (33% to 52%) | 32 | 31% (22% to 41%) | 19 | 18% (11% to 27%) |
| **Bipolar** | 40 | 59 | 9 | 15% (7% to 27%) | 2 | 5% (6% to 17%) | 48 | 81% (69% to 90%) | 21 | 36% (24% to 49%) | 7 | 12% (5% to 23%) | 20 | 34% (22% to 47%) |
| **Anorexia** | 13 | 29 | 4 | 14% (4% to 32%) | 0 | 0 | 25 | 86% (68% to 96%) | 5 | 17% (6% to 36%) | 2 | 7% (8% to 23%) | 18 | 62% (42% to 79%) |
| **Bulimia** | 21 | 45 | 3 | 7% (1% to 18%) | 2 (2) | 4% (0.5% to 15%) | 40 | 89% (76% to 96%) | 1 | 2% (0.5% to 12%) | 2 | 4% (0.5% to 15%) | 37 | 82% (68% to 92%) |
| **Total** | **260** | **422** | **37** | **9% (6% to 12%)** | **21** | **5% (3% to 8%)** | **364** | **86% (83% to 89%)** | **106** | **25% (21% to 30%)** | **67** | **16% (13% to 20%)** | **191** | **45% (40% to 50%)** |

*Note.* AU, Author; BPD, Borderline personality disorder; SUD, Substance Use Disorder; RCT, randomized controlled trial

^a^ Total number of active arms

^b^ Active arms with accessible protocol or manual if it these could be retrieved from any source (published paper, trial registry, author contact, available in library or commercially)

^c^ Calculated out of the total number of arms

^d^ Publicly available protocol or manual, i.e., freely available without restrictions, as for example open-access manuals or publications, preprints, others

^e^ Protocol or manual obtained through author contact

^f^ Protocol or manual only identified as commercially available

^g^ Twelve arms included the same intervention (contingency management) with the only variation being the type of contingency
